# Relation between Methylenetetrahydrofolate Reductase C677T and A1298C Polymorphisms and Migraine Susceptibility

**DOI:** 10.1101/19011601

**Authors:** Vandana Rai, Pradeep Kumar

## Abstract

Migraine is a neurological disorder which impairs the patient’s quality of life. Several association studies investigating the association between MTHFR gene C677T and A1298C polymorphisms and susceptibility to migraine were published. But the results were conflicting, so authors performed a meta-analysis of published case control studies. Four databases were searched for suitable studies up to December, 2018. Odds ratios (OR) with 95% confidence intervals (CI) was calculated adopting additive, homozygote, co-dominant, dominant, and recessive genetic models.

Results of MTHFR C677T polymorphism studies meta-analysis showed significant association with migraine risk using allele contrast, homozygote, dominant and recessive genetic models (T vs. C: OR = 1.18, 95%CI = 1.00-1.26, p= 0.05; TT vs. CC: OR = 1.24, 95%CI = 1.0-1.5, p= 0.04; CT vs. CC: OR = 1.08, 95%CI = 0.97-1.07, p= 0.25; TT+CT vs. CC: OR = 1.15, 95%CI = 1.0-1.29, p= 0.04; TT vs. CT +CC: OR = 1.97, 95%CI = 1.28-3.42, p= 0.002). However, results of MTHFR A1298 polymorphism studies meta-analysis did not show any association with migraine. Subgroup analysis based on ethnicity and migraine types i. e migraine with aura (MA) and without aura (MO) were also performed. Results of present meta-analysis indicate overall association between MTHFR C677T polymorphism with migraine in total 24 studies, in Asian population and in MA cases but did not show any association with Caucasian population and MO cases.

## Introduction

Migraine is a highly prevalent neurological disorder affecting up to 20% of the general population [1]. It is characterized by recurrent episodes of headache, nausea, vomiting, photophobia, phonophobia and autonomic nervous system dysfunction [2]. International Headache Society (IHS) defined two major classes of migraine: migraine with aura (MA) and migraine without aura (MO).

Migraine is considered a polygenic multifactorial disease with several genes participating in its pathogenesis through interaction with environmental factors [3,4]. About 50% of affected individuals have a first-degree relative also suffering from migraine [5-7]. Family and twin studies support the idea of MO and MA being different phenotypes of the same entity, with a heritability ranging from 33 to 57% [6,8,9]. The number and types of genes responsible for migraine are still not clearly known.

Among all the genes associated with common migraine and MA, the 5,10-methylenetetrahydrofolate reductase (MTHFR) gene is the most thoroughly studied one. MTHFR gene is located on chromosome 1 (1p36.3) and contains 11 exons. Several polymorphisms are reported in MTHFR gene, but C677T and A1298C polymorphisms are clinically very important and most studied so far. In C677T polymorphism, cytosine at 677 position is substituted by thymine. This substitution makes MTHFR enzyme thermolabile with reduced enzymatic activity. T allele frequency varied worldwide and well studied in several populations [10-13]. A1298C polymorphism involving alanine to cytosine substitution in MTHFR gene has also been reported to reduce enzyme activity [14]. The prevalence of the A1298C homozygote variant (CC) ranges from 7 to 12% in White populations of North America and Europe. Lower frequencies have been reported in Hispanics (4 to 5 %), and Asian populations (1 to 4%) [15,16]. (The enzyme 5,10-methylenetetrahydrofolate reductase (MTHFR) is an important enzyme in the homocysteine metabolism and catalyzes the conversion of 5,10-methylenetetrahydrofolate into 5-methyltetrahydrofolate, the predominant circulating form of folate. Normal MTHFR activity is crucial to maintain the pool of circulating folate and methionine and to prevent the accumulation of homocysteine [17,18]. The 677T allele is associated with reduced enzyme activity, and mildly increased plasma total homocysteine (tHcy) concentrations [17,18]. MTHFR gene polymorphisms have been reported as risk factor for several neurological and psychiatric diseases/ disorders like-Neural tube defects [19], Down syndrome [20], Alzheimer’s disease [21], Parkinsons’ disease [22], schizophrenia [23], autism [24] and epilepsy [25] etc.

Several case control association studies investigating MTHFR C677T and A1298C polymorphisms role in migraine susceptibility were published but results were contradictory. Some studies reported positive association [26,27] and some other studies showed negative association [28,29]. To clarify the association between C677T and A1298C polymorphisms and migraine risk, present meta-analysis was performed by including more recent publications to improve the efficiency of meta-analysis.

## Methods

Meta-analysis was carried out according to MOOSE guidelines [30].

### Selection of studies

All studies that investigated the association of the MTHFR C677T polymorphism with migraine, published before December, 2018 were considered in the present meta-analysis. These studies were identified by extended computer based search of the PubMed (http://www.ncbi.nlm.nih.gov/pubmed), Google Scholar (http://scholar.google.com), Science Direct (http://www.sciencedirect.com), and Springer Link (http://link.springer.com) databases. The combination of the following terms was used as a search criterion: ‘‘MTHFR’’, C677T’’, “A1298C”,’’methylenetetrahydrofolate reductase”, ‘‘migraine”. All references cited in the retrieved studies were also reviewed to identify additional articles not indexed in these databases.

### Data extraction

The following information about the eligible studies was extracted: first author name, year of publication, country of study, ethnicity of studied subjects, full genotyping data for the case and control groups. The frequencies of the alleles were calculated, for the cases and the controls, from the corresponding genotype distributions.

### Inclusion and exclusion criteria

The inclusion criteria for the present meta-analysis are following: (i) Studies must have case-control or cohort design. (ii) Authors must investigated patients with migraine and healthy control subjects. (iii) Authors must provided information on genotype numbers/frequencies or sufficient data to calculate these. (iv) Studies must be published as full articles.

Studies were excluded if: 1) they were case reports, editorials review, letter to editors and book chapters (2) incomplete raw data/information and not providing complete information for number of genotype and/or allele number calculation, 3) studies based on pedigree, and genome scans, since they investigate linkage.

### Statistical analysis

The meta-analysis examined the overall association of T (C677T polymorphism) and C (A1298C polymorphism) alleles and risk of migraine compared with T and A alleles respectively, using the allele contrast/additive model, homozygote model, co-dominant/heterozygote model, recessive model and dominant model. The effect of association was indicated as odds ratio (OR) with the corresponding 95% confidence interval (CI). The pooled OR was estimated using fixed effects (FE) [31] and random effects (RE) model [32]. RE modeling assumes heterogeneity between the studies, and it incorporates the between-study variability. The heterogeneity between studies was tested by the Q statistic [33]. If p<0.05 then the heterogeneity was considered statistically significant. Heterogeneity was quantified by the I^2^ metric (I^2^<25% no heterogeneity; I^2^=25–50% moderate heterogeneity; I^2^>50% large or extreme heterogeneity) [34]. The distribution of the genotypes in the control group was tested for Hardy-Weinberg equilibrium using calculator available at http://ihg.gsf.de/cgi-bin/hw/hwa1.pl. Studies with the controls not in Hardy-Weinberg equilibrium (HWE) were subjected to a sensitivity analysis [35], i.e., the effect of excluding specific studies was examined. Subgroup analysis based on ethnicity was also performed.

Publication bias was tested by funnel plot asymmetry using Egger’s linear regression test. The significance of the intercept was determined by the t-test considering p-value < 0.05 as representation of statistically significant publication bias [36]. All analyses were performed using the computer programs MetaAnalyst [37] and MIX version 1.7 [38]. A p value less than 0.05 was considered statistically significant, and all the p values were two sided.

## Results

### Eligible studies

A diagram schematizing the study selection process is presented in Figure 1. Initial PubMed, Google Scholar, Science Direct and Elsevier Link databases search, total 126 studies were retrieved. After title and abstract evaluation, 83 articles were eliminate, which were irrelevant for the present meta-analysis. These 83 publications were reviews, case studies, editorials, comments, reviews, meta-analysis etc. Following inclusion and exclusion criteria, total 24 case–control studies were found suitable for the meta-analysis [26-29,39-58] (Table 1). Out of 24 studies, only in three studies A1298C polymorphism was investigated (Table 2).

**Table 1.** Details of MTHFR C677T genotypes in twenty four included studies

| Study | Country | Control/Case | Genotypes in cases<br>CC/CT/TT | Genotypes in controls<br>CC/CT/TT | HWE<br>p-value |
| --- | --- | --- | --- | --- | --- |
| Kowa et al.,2000 | Japan | 261/74 | 18/41/15 | 104/132/25 | 0.06 |
| Kara et al.,2003 | Turkey | 136/93 | 36/49/8 | 69/65/2 | 0.002 |
| Lea et al.,2004 | Australia | 269/268 | 104/125/39 | 117/129/23 | 0.12 |
| Oterino et al.,2004 | Spain | 204/230 | 105/98/27 | 84/93/27 | 0.87 |
| Oterino et al.,2005 | Italy | 237/329 | 142/147/40 | 94/114/29 | 0.53 |
| Scher, et al.,2006 | Netherlands | 1212/413 | 181/186/46 | 567/527/118 | 0.78 |
| Todt et al.,2006 | Germany | 625/656 | 300/279/77 | 251/300/74 | 0.27 |
| Kaunisto et al.,2006 | Finland | 900/898 | 521/332/45 | 522/324/54 | 0.69 |
| Botini et al.,2006 | Italy | 66/45 | 16/17/12/ | 24/33/9 | 0.65 |
| Pezzini et al.,2007 | Italy | 105/206 | 75/90/41 | 41/51/13 | 0.21 |
| Schurks et al.,2008 | USA | 20424/4577 | 2070/2038/469 | 9173/8939/2312 | 0.57 |
| Ferro et al.,2008 | Portugal | 96/186 | 79/91/16 | 35/47/14 | 0.78 |
| Joshi et al.,2009 | India | 150/150 | 54/78/18 | 60/78/12 | 0.05 |
| Oterino et al.,2010 | Spain | 310/427 | 183/184/60 | 117/147/46 | 0.98 |
| Schurks et al.,2010 | Iceland | 1357/252 | 116/114/22 | 612/579/166 | 0.11 |
| Samaan et al.,2011 | Canada | 1402/447 | 181/204/62 | 625/596/181 | 0.05 |
| Ishii et al.,2012 | Japan | 119/91 | 42/31/18 | 37/61/21 | 0.63 |
| An et al.,2012 | China | 137/151 | 67/60/24 | 80/46/11 | 0.24 |
| Azimova et al. ,2013 | Russia | 50/83 | 32/33/18 | 26/20/4 | 0.95 |
| Bahadir et al.,2013 | Turkey | 107/150 | 50/59/41 | 96/10/1 | 0.22 |
| Scher et al.,2013 | USA | 1357/252 | 116/114/22 | 612/579/166 | 0.11 |
| Essmeister et al.,2016 | Austria | 420/244 | 189/215/16 | 114/121/9 | 0.000 |
| Kaur et al.,2018 | India | 23/100 | 6/14/3 | 51/42/7 | 0.67 |
| Salehi et al.,2018 | Iran | 223/275 | 107/91/25 | 159/101/15 | 0.84 |

**Table 2.** Details of MTHFR A1298C genotypes in three included studies

| Study | Country | Control/Case | Genotypes in cases<br>AA/AC/CC | Genotypes in controls<br>AA/AC/CC | HWE<br>p-value |
| --- | --- | --- | --- | --- | --- |
| Kara et al.,2003 | Turkey | 136/102 | 45/45/12 | 72/62/2 | 0.005 |
| Kaur et al.,2018 | India | 77/23 | 6/11/6 | 23/30/24 | 0.05 |
| Salehi et al.,2018 | Iran | 275/223 | 81/101/41 | 100/134/41 | 0.72 |

**Figure 1.**
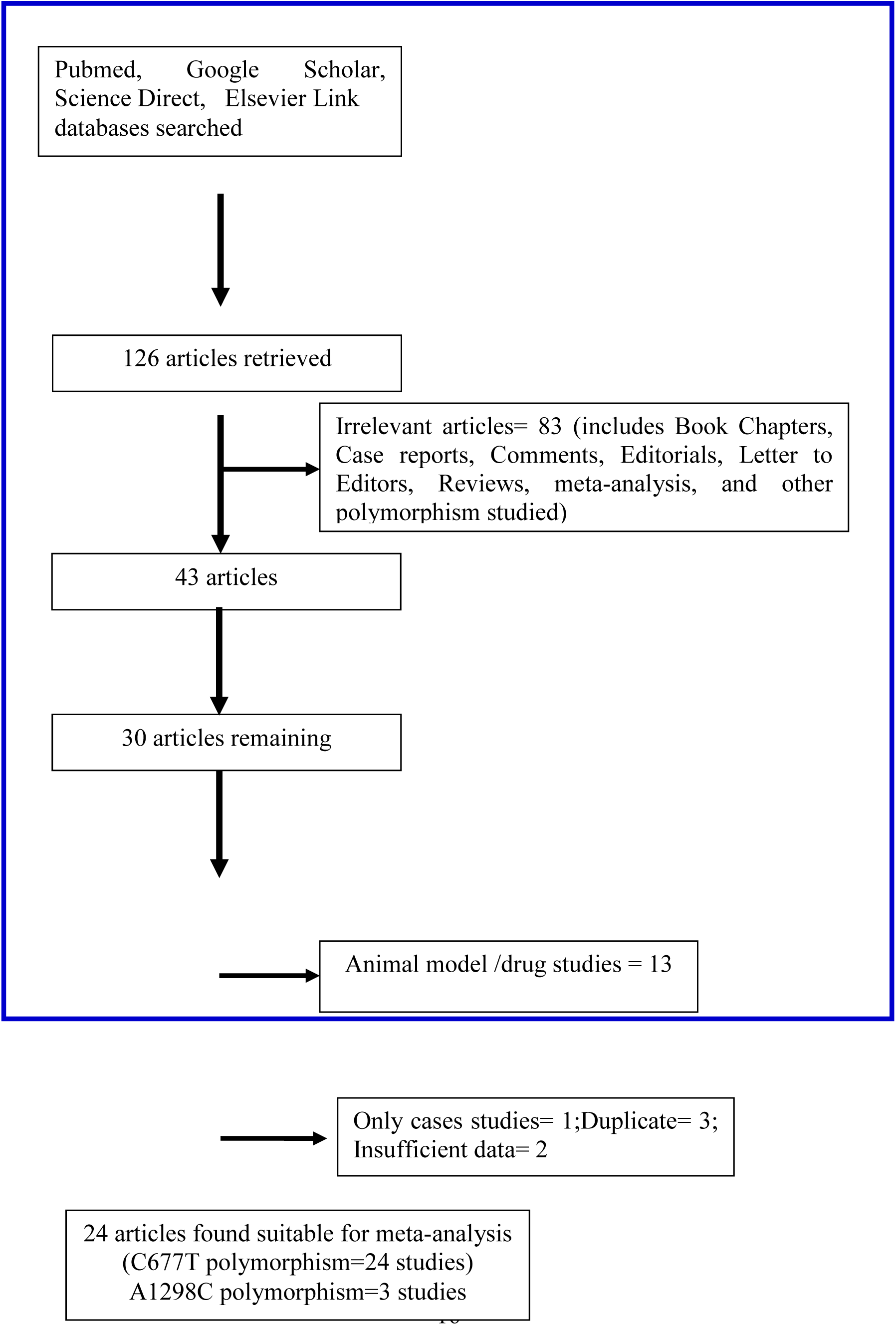
Flow Diagram of Study Search and Selection Process.

These studies were reported from different countries like-Australia [41], Austria [56], Spain [42], Netherlands [44], Germany [45], Finland [28], Iceland [52], India [50,57], Iran [58], Italy [43,46,47], Portugal [49], Canada [53], Japan [29,39], China [54], Russia [26], Spain [51], Turkey [27,40], and USA [48,55] (Table 1).

### Summary statistics

In MTHFR C677T polymorphism studies, number of cases and controls were 10,644 and 30,143 respectively. Except two studies [40,56], distribution of the C677T genotype in the control group of all 22 studies was in Hardy–Weinberg equilibrium (p<0.05), indicating a lack of genotyping, sampling errors and/or population stratification [59] (Table 1). Odds ratio in ten studies was below one and did not reported association[28,29,42,43,45,48,49,51,52,55], and other fourteen studies reported association between MTHFR C677T polymorphism and migraine. In cases, the frequency of CC, CT and TT genotypes were 45.0 %, 44.1 % and 11 % respectively. In control groups, the frequencies of CC, CT and TT-homozygous individuals were 45.35%, 43.57%, and 11.07 %, respectively.

In MTHFR A1298C polymorphism studies, number of cases and controls were 348 and 488 respectively. Except Kara et al.[40], distribution of the A1298C genotype in the control group of all 2 studies was in Hardy–Weinberg equilibrium (p<0.05). In cases, the frequency of AA, AC and CC genotypes were 37.93 %, 45.11 % and 16.95% respectively. In control groups, the frequencies of AA, AC and CC genotypes were 39.95 %, 46.31 % and 13.72% respectively.

### MTHFR C677T meta-analysis

Meta-analysis of 24 studies for investigating the association of the MTHFR C677T polymorphism, showed higher heterogeneity using all five genetic models (p<0.0001, I^2^= 87.57%); so random effect model was adopted. The random effects pooled OR was not statistically significant (T vs C: OR= 1.18; 95% CI=1.00-1.26; p=0.05; CT vs CC: OR= 1.08; 95% CI = 0.97-1.07; p=0.25; TT vs CC: OR= 1.24; 95% CI = 1.0-1.51; p=0.04; TT+CT vs CC: OR= 1.15; 95% CI = 1.0-1.29; p=0.04; TT vs CT+CC: OR= 1.97; 95% CI = 1.28-3.42; p=0.002) (Table 3; Figure 2).

**Table 3:** Summary estimates for the odds ratio (OR) of MTHFR C677T in various allele/genotype contrasts, the significance level (p value) of heterogeneity test (Q test), and the I^2^ metric and publication bias p-value (Egger Test).

| Genetic Models | Fixed effect<br>OR (95% CI), p | Random effect<br>OR (95% CI), p | Heterogeneity p-value (Q test) | $I^2$ (%) | Publication Bias (p of Egger's test) |
| --- | --- | --- | --- | --- | --- |
| Allele Contrast (T vs C) | 1.00(0.99-1.02),0.64 | 1.18(1.00-1.26),0.05 | <0.0001 | 87.57 | 0.099 |
| Co-dominant (Ct vs CC) | 1.02(0.97-1.07),0.36 | 1.08(0.97-1.07),0.25 | <0.0001 | 70.0 | 0.39 |
| Homozygote (TT vs CC) | 1.20(0.99-1.1),0.91 | 1.24(1.0-1.51),0.04 | <0.0001 | 70.86 | 0.06 |
| Dominant (TT+CT vs CC) | 1.03(0.98-1.08),0.24 | 1.15(1.00-1.29),0.04 | <0.0001 | 81.2 | 0.12 |
| Recessive (TT vs CT+CC) | 1.01(0.64-0.73),<0.0001 | 1.97(1.28-3.42),0.002 | <0.0001 | 95.68 | <0.001 |
| **Asian studies (study)** |  |  |  |  |  |
| Allele Contrast (T vs C) | 1.90(1.13-1.58),0.008 | 1.91 (1.06-3.45),0.03 | 0.01 | 67.4% | 0.78 |
| Co-dominant (Ct vs CC) | 1.55(1.24-1.98),<0.0001 | 1.47(1.16-1.85),0.001 | <0.0001 | 88.76 | 0.28 |
| Homozygote (TT vs CC) | 2.99(2.13-4.22),<0.001 | 2.98(1.56-5.68),0.008 | <0.0001 | 80.79 | 0.054 |
| Dominant (TT+CT vs CC) | 1.84(1.49-2.26),<0.0001 | 1.96(0.90-4.24),0.08 | <0.0001 | 91.81 | 0.192 |
| Recessive (TT vs CT+CC) | 2.58(1.86-3.5),<0.0001 | 2.5(1.33-4.67),0.004 | 0.008 | 67.57 | 0.02 |
| **Caucasian studies (study)** |  |  |  |  |  |
| Allele Contrast (T vs C) | 0.94(0.93-1.01),0.19 | 0.97(0.87-1.08),0.19 | <0.0001 | 90.11% | 0.02 |
| Co-dominant (Ct vs CC) | 0.99(0.94-1.05),0.99 | 0.99(0.95-1.05),0.99 | 0.58 | 0 | 0.44 |
| Homozygote (TT vs CC) | 0.94(0.86-1.01),0.12 | 0.99(0.86-1.15),0.95 | 0.02 | 45.31 | 0.13 |
| Dominant (TT+CT vs CC) | 0.98(0.94-1.03),0.59 | 0.98(0.92-1.05),0.66 | 0.33 | 10.37 | 0.94 |
| Recessive (TT vs CT+CC) | 0.93(0.86-1.02),0.12 | 0.99(0.86-1.15),0.95 | 0.02 | 45.31 | 0.14 |
| **Migraine with Aura (study)** |  |  |  |  |  |
| Allele Contrast (T vs C) | 1.11(1.03-1.18),0.003 | 1.26(1.03-1.54),0.02 | <0.0001 | 87.09 | 0.09 |
| Co-dominant (Ct vs CC) | 1.03(0.93-1.13),0.59 | 1.1(0.87-1.37),0.40 | <0.0001 | 73.29 | 0.14 |
| Homozygote (TT vs CC) | 1.31(1.12-1.52),0.0005 | 1.51(1.05-2.17),0.02 | <0.0001 | 76.09 | 0.148 |
| Dominant (TT+CT vs CC) | 1.08(0.99-1.18),0.09 | 1.21(0.94-1.55),0.14 | <0.0001 | 82.22 | 0.11 |
| Recessive (TT vs CT+CC) | 1.3(1.13-1.5),0.0003 | 1.45(1.06-1.96),0.02 | <0.0001 | 71.18 | 0.28 |
| **Migraine without aura (study)** |  |  |  |  |  |
| Allele Contrast (T vs C) | 0.99(0.91-1.07),0.80 | 1.07(0.84-1.36),0.56 | <0.0001 | 87.5 | 0.22 |
| Co-dominant (Ct vs CC) | 0.99(0.88-1.10),0.87 | 1.02(0.78-1.32),0.88 | <0.0001 | 78.58 | 0.64 |
| Homozygote (TT vs CC) | 0.95(0.80-1.14),0.62 | 1.05(0.70-1.56),0.81 | <0.0001 | 72.89 | 0.17 |
| Dominant (TT+CT vs CC) | 0.99(0.89-1.10),0.94 | 1.06(0.78-1.42),0.7 | <0.0001 | 85.2 | 0.40 |
| Recessive (TT vs CT+CC) | 0.96(0.81-1.14),0.69 | 1.02(0.74-1.41),0.88 | 0.0004 | 62.68 | 0.11 |

**Figure 2.**
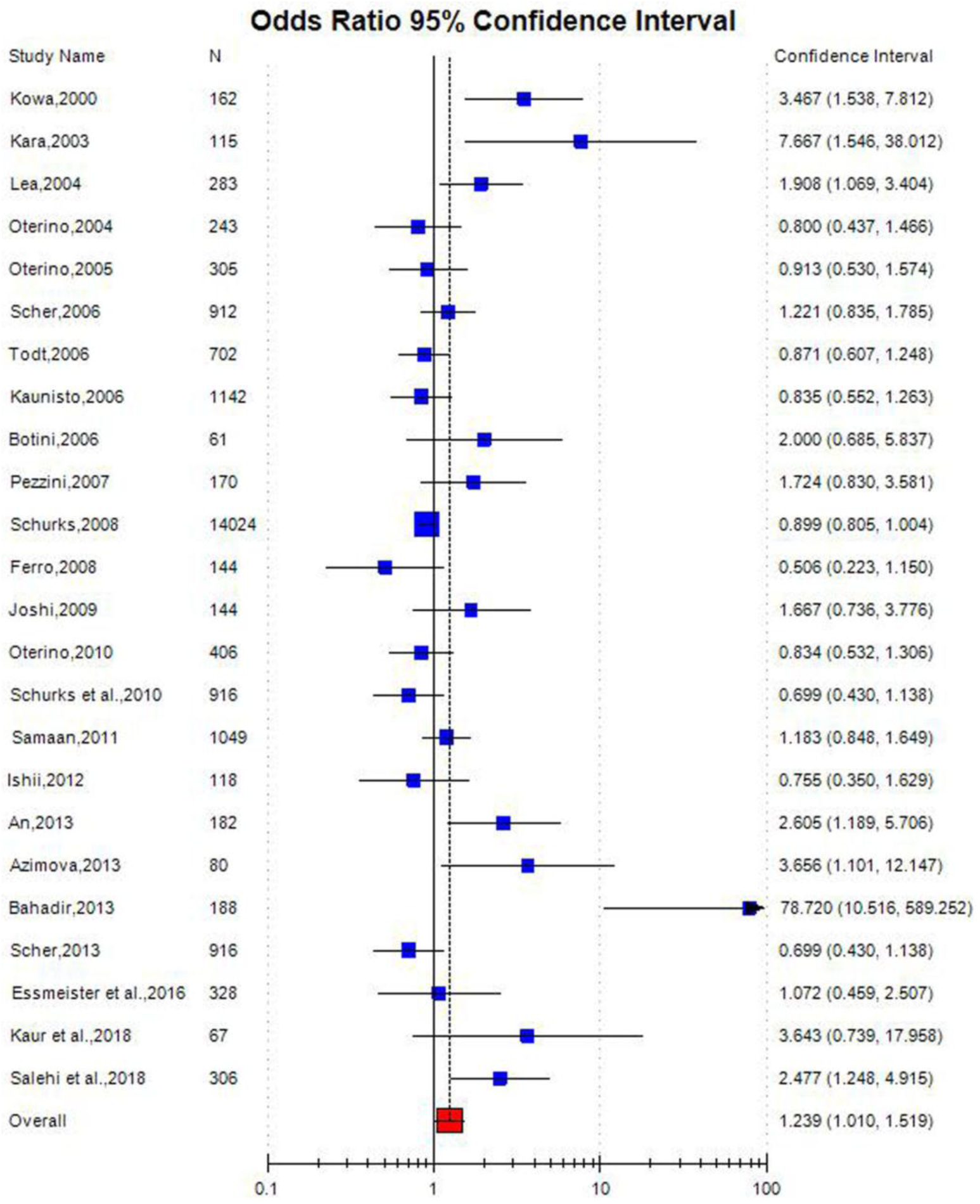
Random effect Forest plot of MTHFR C677T polymorphism of homozygote model (TT vs. CC) of total 24 studies.

### MTHFR A1298C meta-analysis

Meta-analysis of three A1298C studies showed that MTHFR A1298C polymorphism was not associated with migraine (C vs A: OR = 1.18, 95% CI = 0.96–1.44, p=0.11; AC vs AA: OR = 1.03, 95% CI = 0.76–1.39, p=0.84; CC vs AA: OR = 1.55, 95% CI = 1.0–2.4, p=0.05; CC+AC vs AA: OR = 1.13, 95% CI = 0.85–1.51, p=0.39; CC vs AC+AA: OR = 1.47, 95% CI = 0.98–2.19, p=0.05) (Table 4, Figure 3).

**Table 4:** Summary estimates for the odds ratio (OR) of MTHFR A1298C in various allele/genotype contrasts, the significance level (p value) of heterogeneity test (Q test), and the I^2^ metric and publication bias p-value (Egger Test).

| **Genetic Models** | **Fixed effect**<br>**OR (95% CI), p** | **Random effect**<br>**OR (95% CI), p** | **Heterogeneity**<br>**p-value (Q**<br>**test)** | **$I^2$ (%)** | **Publication**<br>**Bias (p of**<br>**Egger's test)** |
| --- | --- | --- | --- | --- | --- |
| Allele Contrast (T vs C) | 1.18(0.96-1.44),0.11 | 1.2(0.90-1.58)0.19 | 0.22 | 33.36 | 0.88 |
| Co-dominant (Ct vs CC) | 1.03(0.76-1.39),0.84 | 1.02(0.75-1.39),0.85 | 0.69 | 0 | 0.32 |
| Homozygote (TT vs CC) | 1.55(1.0-2.4)0.05 | 1.9(0.62-5.98),0.25 | 0.03 | 69.97 | 0.57 |
| Dominant (TT+CT vs CC) | 1.13(0.85-1.51),0.39 | 1.13(0.85-1.51),0.39 | 0.54 | 0 | 0.68 |
| Recessive (TT vs CT+CC) | 1.47(0.98-2.19),0.05 | 1.7(0.61-4.91),0.30 | 0.02 | 71.92 | 0.04 |

**Figure 3.**
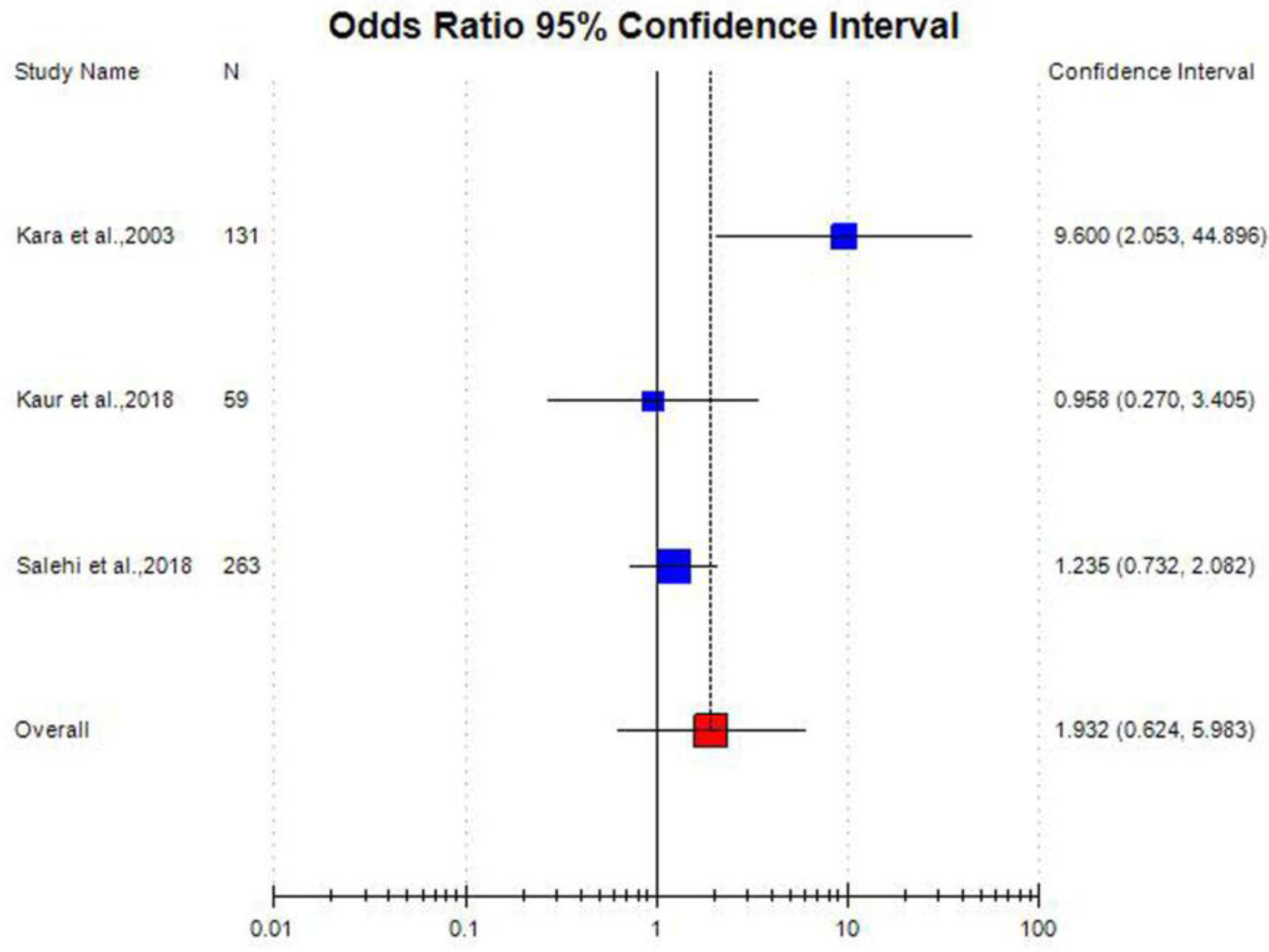
Random effect Forest plot of MTHFR A1298C polymorphism of homozygote model (CC vs. AA) of 3 Asian studies.

### Subgroup analysis

Subgroup analysis were performed on the bases of ethnicity and type of migraine i. e MA and MO. In total 24 studies, Asian and Caucasian subjects were genotyped in eight and sixteen studies respectively. Sixteen Caucasian studies meta-analysis included 9,689 cases and 28,858 controls did not show association between C677T and migraine risk (T vs C: OR= 0.97; 95% CI=0.93-1.01; p=0.19; TT vs CC: OR= 0.99; 95% CI=0.86-1.15; p=0.95) (Table 3, Figure 4). Meta-analysis of 8 Asian studies (955 cases and 1,285 controls) showed strong significant association (T vs C: OR= 1.91; 95% CI=1.06-3.45; p=0.03; TT vs. CC: OR= 2.98; 95% CI=1.56-5.68; p=0.008) between C677T polymorphism and migraine risk (Table 3, Figure 5).

**Figure 4.**
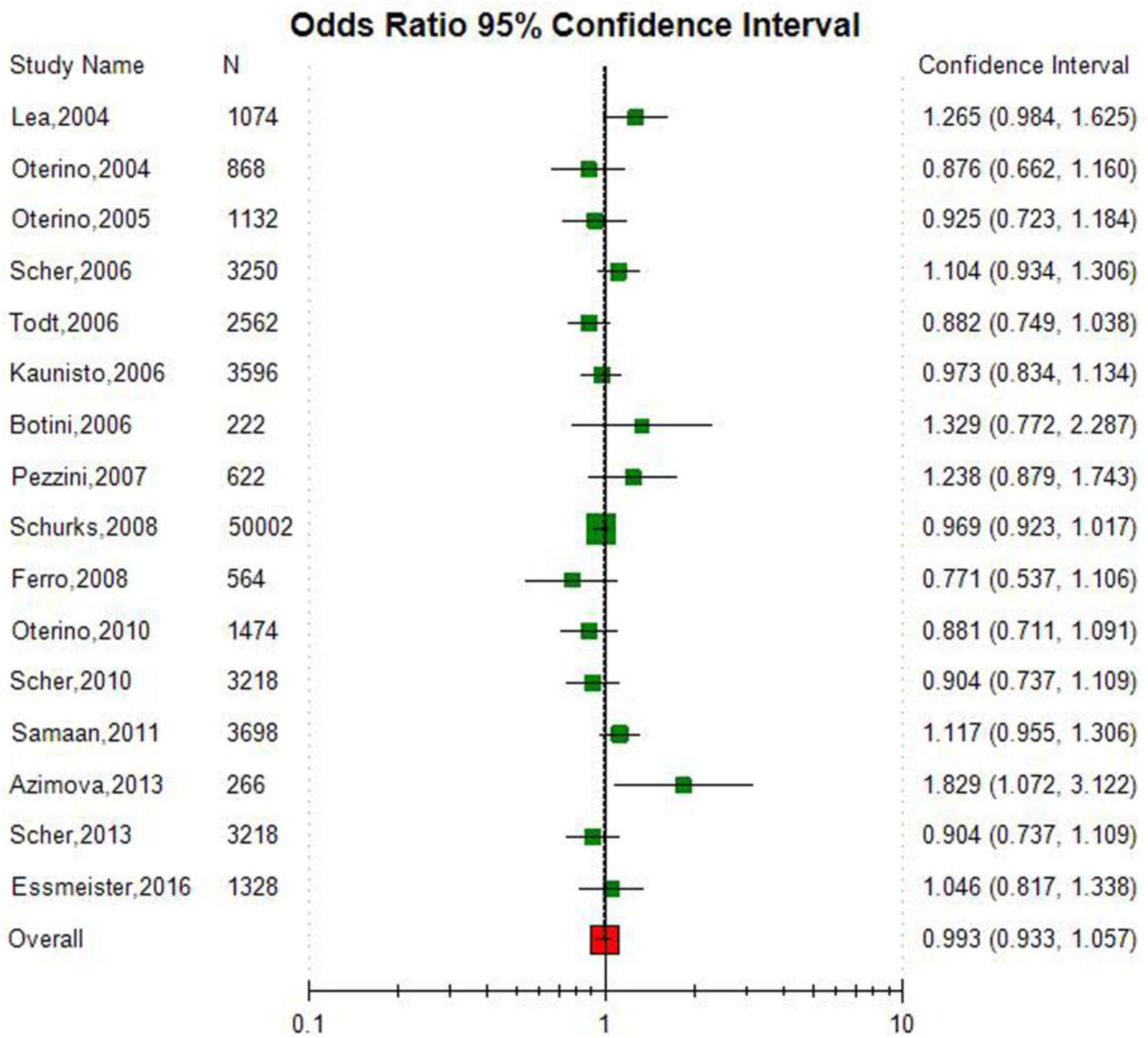
Random effect Forest plot of MTHFR C677T polymorphism of allele contrast model (T vs. C) of 16 Caucasian studies.

**Figure 5.**
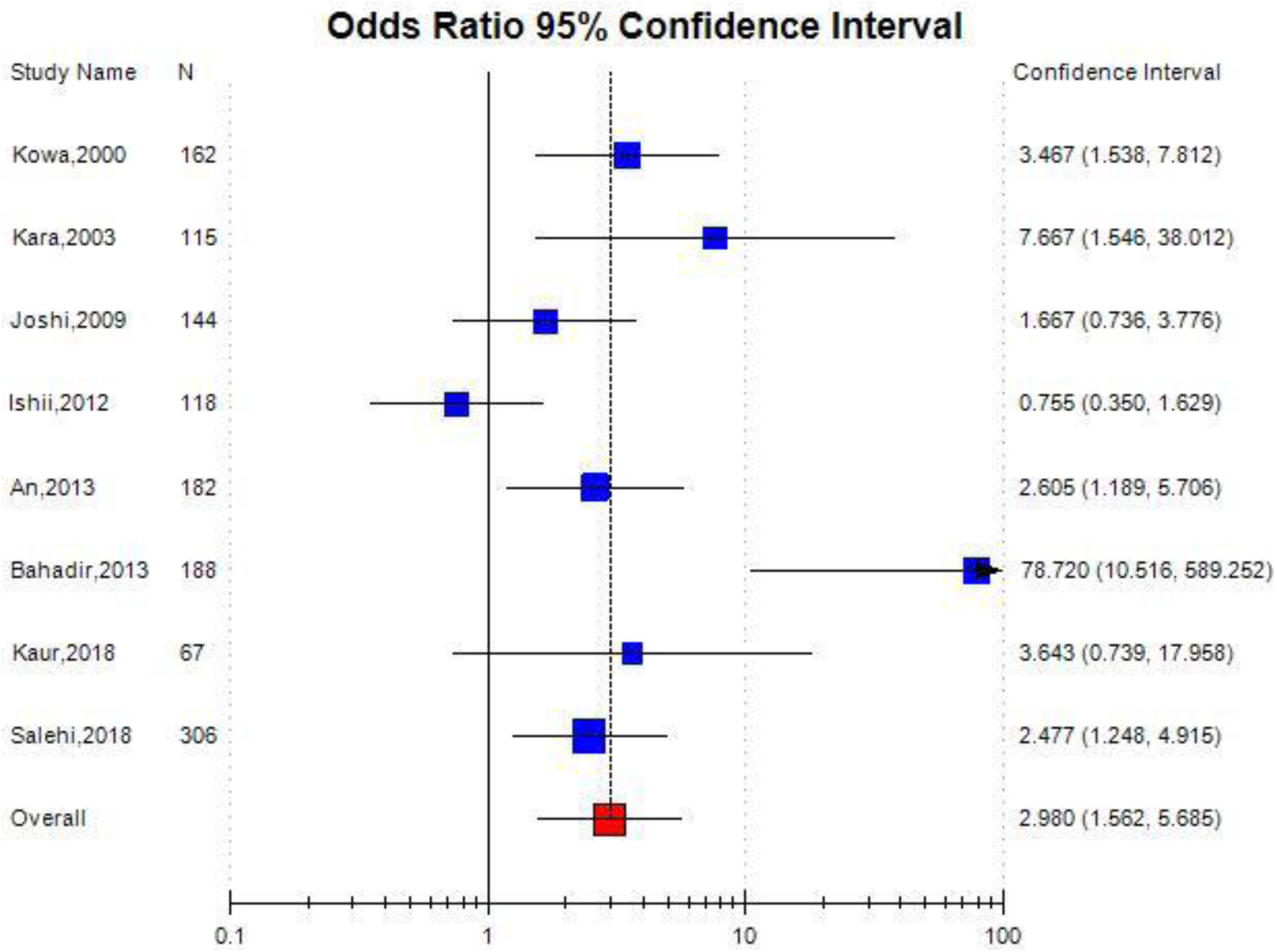
Random effect Forest plot of MTHFR C677T polymorphism of homozygote model (TT vs. CC) of 8 Asian studies.

Out of total 24 studies, 19 studies included 3,177 MA cases and 7,921 controls. Allele contrast meta-analysis of MA studies showed significant association between C677T polymorphism and MA (T vs C: OR= 1.26; 95%CI= 1.03-1.54; p=0.02; TT vs CC: OR= 1.51; 95%CI= 1.05-2.17; p=0.02) (Table 3, Figure 6). Higher significant heterogeneity was observed but publication bias was not observed in meta-analysis of MA studies. Meta-analysis of seventeen MO studies with 2347 cases and 6406 controls, did not show association between C677T polymorphism and MO (OR= 1.07;95%CI= 0.84-1.36; p= 0.56) (Table 3, Figure 6).

**Figure 6:**
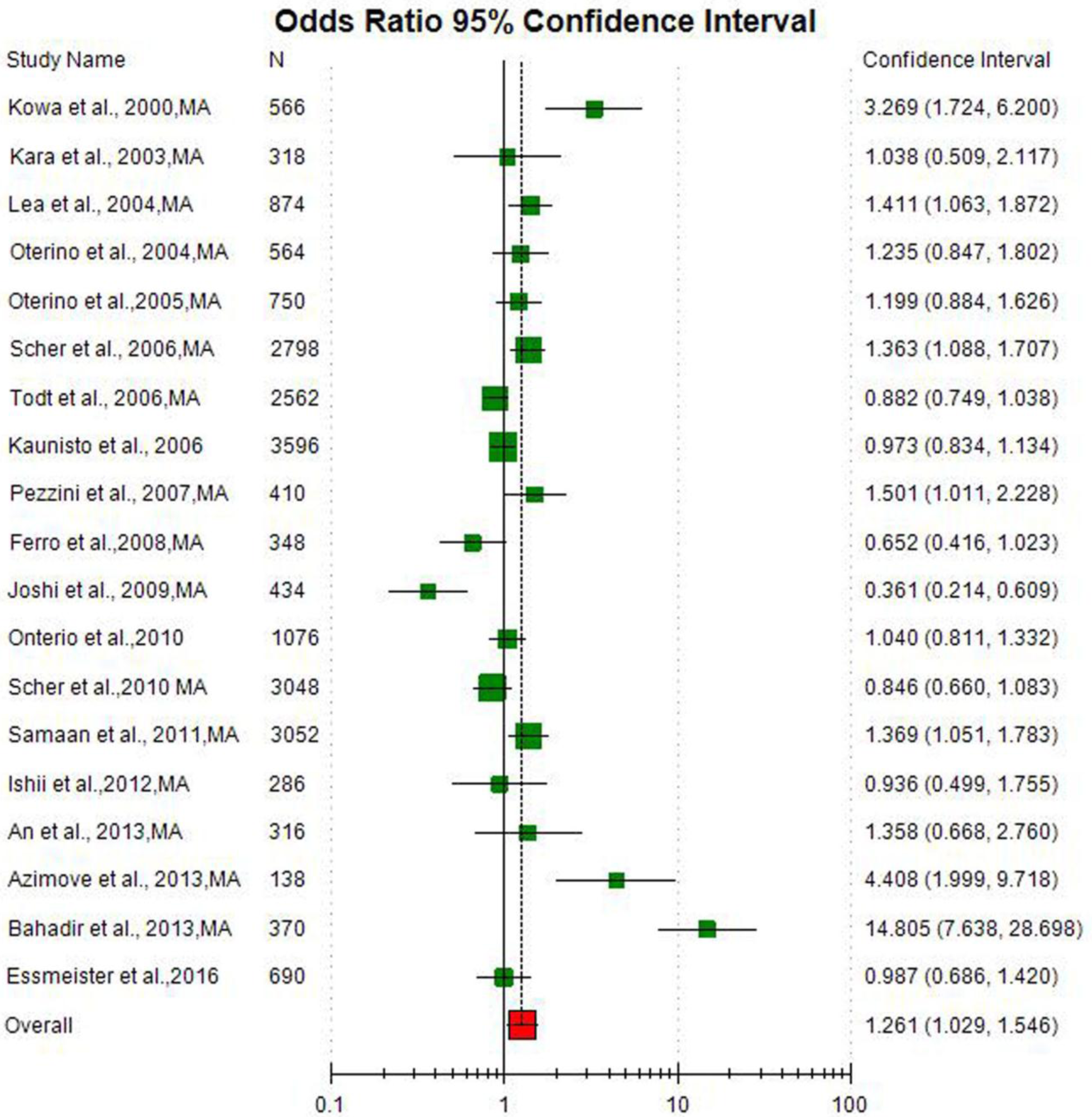
Random effect Forest plot of MTHFR C677T polymorphism of allele contrast model (T vs. C) of 19 studies of migraine with aura cases.

**Figure 7.**
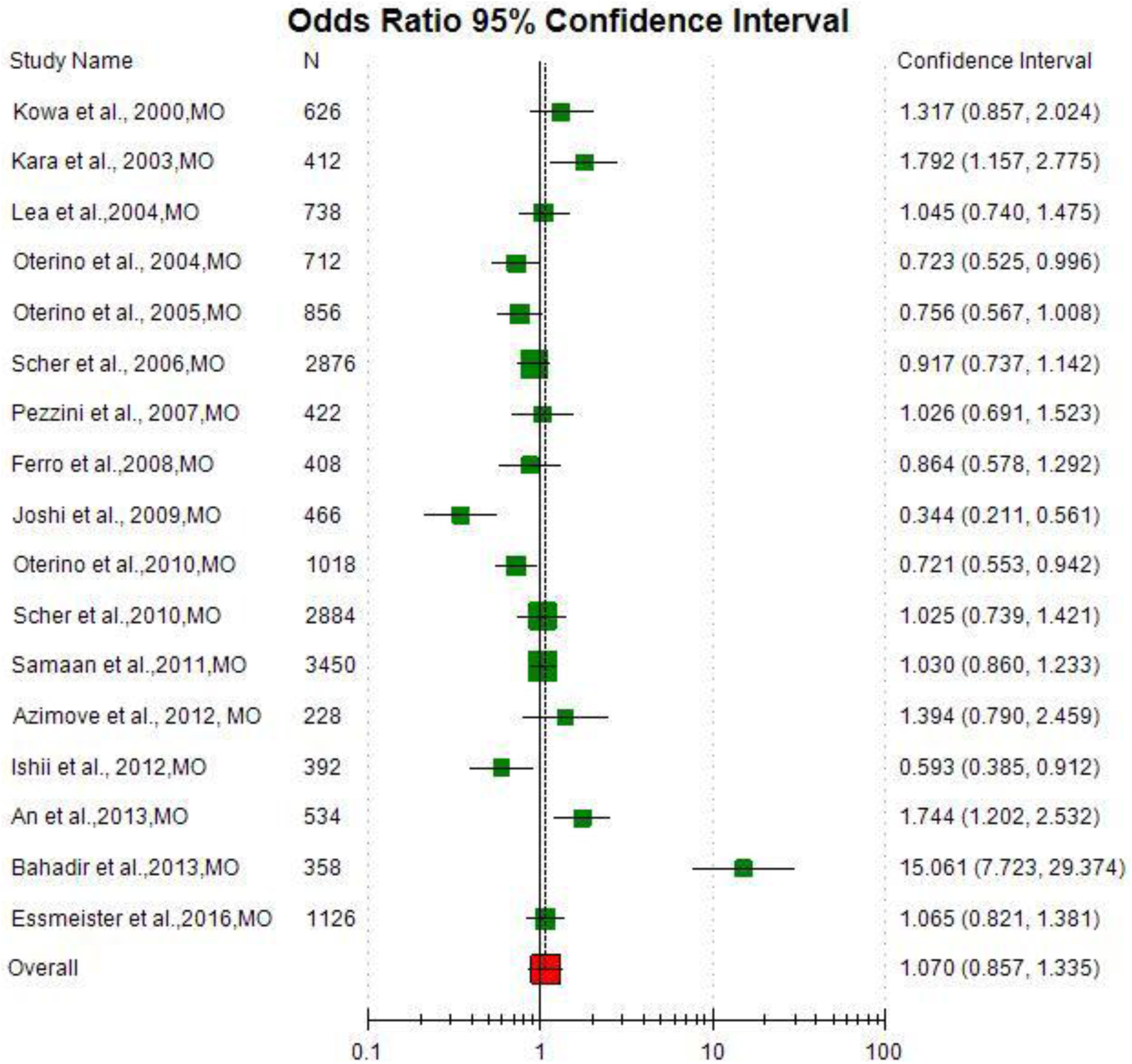
Random effect Forest plot of MTHFR C677T polymorphism of allele contrast model (T vs. C) of 17 studies of migraine without aura cases.

### Publication bias

In total MTHFR C677T studies meta-analysis, except recessive model publication bias was not observed in other four models (Egger’s p= 0.09 for T vs. C; Egger’s p= 0.06 for TT vs. CC; Egger’s p= 0.39 for CT vs. CC; Egger’s p= 0.12 for TT + CC vs. CT; Egger’s TT vs. CT+CC<0.0001) of overall meta-analysis (Table 3,Figure 8). In A1298C studies, publication bias was absent (Table 4).

**Figure 8.**
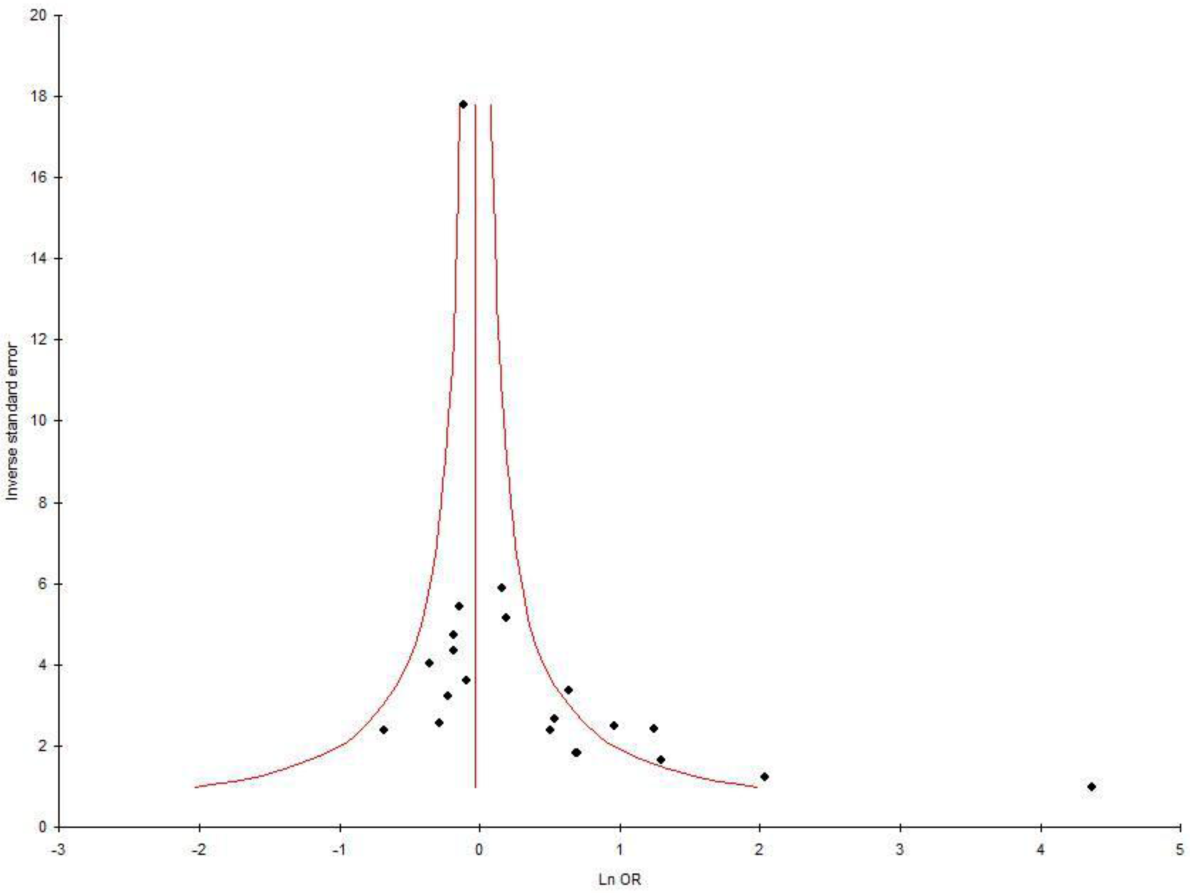
Funnel plot - Precision by log odds ratio for allele contrast model of total 24 studies.

## Discussion

Present meta-analysis was based on data from 24 case-control studies with 10,644 migraine cases and 30,143 controls. Results showed significant association between C677T polymorphism and migraine risk in total 24 studies, Asian populations and MA cases. A1298C polymorphism meta-analysis did not show any association with migraine risk.

Although patho-physiology of migraine is not very well known, but it is considered as disorder of brain with endothelial dysfunction [60] and altered vascular reactivity [61]. TT genotype and/or T allele may lead moderate hyperhomocyeteinemia [40,41] and it is well established fact that higher concentration of homocysteine is toxic to neurons and causes DNA stand breaks, and altered DNA repair, DNA methylation and oxidative stress [62,63,64,65]. Hyperhomocysteinemia produce endothelial cell injury, which may activate trigeminovascular system (TVS), resulting in an inflammatory action in the meninges and dilation of the large cerebral vessels. These changes may start the progression of migraine [40,41,66]. The characteristic head pain in migraine may arise due to dilation of cerebral blood vessels following activation of the TVS [41].

Meta-analysis is a useful strategy for elucidating genetic factors in different diseases/disorders. Several meta-analysis were published which evaluated risk of folate pathway genes polymorphism for different disease and disorders-like Down syndrome [67,68], glucose 6-phosphate dehydrogenase deficiency [69], cleft lip/palate [70], neural tube defects [71], recurrent pregnancy loss [72], endometrial cancer [73], male infertility [74], depression [75], schizophrenia [76,77], epilepsy [78], autism [79], Alzheimer’s disease [80], prostate cancer [81], breast cancer [82,83], colorectal cancer [84], Esophageal cancer [85], and digestive tract cancer [86] etc.

Four meta-analyses were published so far in order to draw a reliable conclusion regarding association between C677T polymorphism and migraine susceptibility [52,53,87,88]. Rubino et al. [87] conducted a meta-analysis associating the C677T polymorphism with migraine based on nine published articles (2961 migraineurs, 2170 with MA and 791 with MO), providing evidence for an association of the MTHFR gene only in MA (FE: OR =1.30, 95% CI= 1.06-1.58; RE: OR= 1.66, 95% CI= 1.06-2.59).

Similar observation that the MTHFR 677TT genotype is associated with an increased risk for MA among non-Caucasian population (OR = 1.48, 95% CI= 1.02-2.13) was supported by the meta-analysis of 13 studies by Schurks and co-workers [52]. On the contrary, Samaan et al. [53] including five datasets of Caucasians demonstrated that the TT genotype was associated with total migraine in non-Caucasian population, whereas for Caucasians, this variant was associated with MA only (OR 1.31, 95% CI 1.01-1.70, p = 0.039). A meta-analysis of 16 case-control studies showed that T allele homozygosity is significantly associated with MA (OR = 1.42; 95% CI, 1.10-1.82) and total migraine (OR = 1.37; 95% CI, 1.07-1.76), but not migraine without aura (OR = 1.16; 95% CI, 0.36-3.76) [88]. In present meta-analysis largest number of studies and largest number of samples (40,787) were included.

Similar to other meta-analysis, present meta-analysis have also few limitations, which should be acknowledged like- (i) the pooled OR was based on unadjusted individual ORs, (ii)higher between studies heterogeneity was present, although author tried to eliminate that by performing subgroup analysis, (iii) publication bias was present, which denoted that the some studies with negative results were not included in the present meta-analysis, (iv) owing to lack of information, gene-gene and gene environment interactions could not be carried out in present meta-analysis. Along with limitations, present meta-analysis had several strengths also like (i) highest number of studies and samples were included (ii) controls were healthy and were matched to cases and (iii) subgroup analysis was performed.

In conclusion, the results of present meta-analysis showed significant association between migraine risk with MTHFR C677T polymorphism (OR= 1.24) but did not show any association with MTHFR A1298C polymorphism. In ethnicity based subgroup analysis, positive significant association was found between C677T polymorphism and Asian migraine cases (OR= 2.98) but not with Caucasian cases (OR= 0.99). Further migraine subgroup meta-analysis showed association between C677T polymorphism and migraineurs with aura (OR= 1.51) but not with migraineurs without aura.

## Data Availability

All data included in the manuscript

## Ethical Approval

Present manuscript is a meta-analysis/review, hence, ethical clearance is not required for the present manuscript, no human blood/ tissue samples are used in the present manuscript.

